# Excess ventilation and chemosensitivity in patients with chronic coronary syndrome and patients with heart failure with reduced ejection fraction – a case control study

**DOI:** 10.1101/2024.08.08.24311710

**Authors:** Eser Prisca, Kaesermann Dominic, Calamai Pietro, Kalberer Anja, Stuetz Laura, Huber Sarina, Duffin James, Wilhelm Matthias

## Abstract

**Background:** In patients with chronic coronary syndromes (CCS) increased ventilation/carbon dioxide production (V̇_E_/V̇CO_2_) slope has been found to predict disease progression and mortality similarly to patients with heart failure (HF), however, chemosensitivity has rarely been assessed in patients with CCS.

**Method:** Patients with CCS, HF with reduced ejection fraction (EF<50%), old healthy (45+ years) and young adult healthy controls (<35 years) were recruited. For patients, a V̇_E_/V̇CO_2_ slope ≥36 was an inclusion criterion. The Duffin rebreathing method was used to determine the resting end-expiratory partial pressure of carbon dioxide (P_ET_CO_2_), ventilatory recruitment threshold (VRT) and slope (sensitivity) during a hyperoxic (150 mmHg O_2_) and hypoxic (50 mmHg O_2_) rebreathing test to determine central and peripheral chemosensitivity.

**Results:** In patients with CCS, HF, and old and young controls, median V̇_E_/V̇CO_2_ slopes were 40.2, 41.3, 30.5 and 28.0, respectively. Both patient groups had similarly reduced hyperoxic VRT (at P_ET_CO_2_ 42.1 and 43.2 mmHg) compared to 46.0 and 48.8 mmHg in the old and young controls. Neither hypoxic VRT nor hyper- or hypoxic slopes were significantly different in patients compared to controls. Both patient groups had lower resting P_ET_CO_2_ than controls, but only patients with HF had increased breathing frequency and rapid shallow breathing at rest.

**Conclusion:** In patients with cardiac disease and excess ventilation, central chemoreflex VRT was reduced independently of the presence of heart failure. Low VRTs were related to resting excess ventilation in patients with CCS or HF, however, rapid shallow breathing was present only in patients with HF.

**Clinical perspective:** *What is new?:*

- Excess ventilation during exercise and heightened chemosensory reflexes may be present not only in patients with HF but also in patients with CCS. This suggests that there is a gradual derangement of neurologic and/or hormonal factors leading to excess ventilation before the establishment of HF.
- In patients with excess ventilation during exercise there is also excess ventilation at rest.
- Excess ventilation in patients with CCS does not show the rapid shallow breathing pattern that is typical for patients with HF.

*What are the clinical implications?:*

- While excess ventilation during exercise causes dyspnoea with associated negative effects on exercise tolerance and quality of life,^1^ excess ventilation at rest has been poorly investigated. More research is warranted as physiologic consequences may be substantial with the large time spent at rest compared to exercise.
- The finding that the threshold of P_ET_CO_2_ at which ventilation starts to increase rather than the V̇_E_/P_ET_CO_2_ slope is increased in patients with inefficient ventilation suggests electrolyte derangement as an at least contributing cause which may stimulate alternative treatments such as intravenous iron therapy.^2^

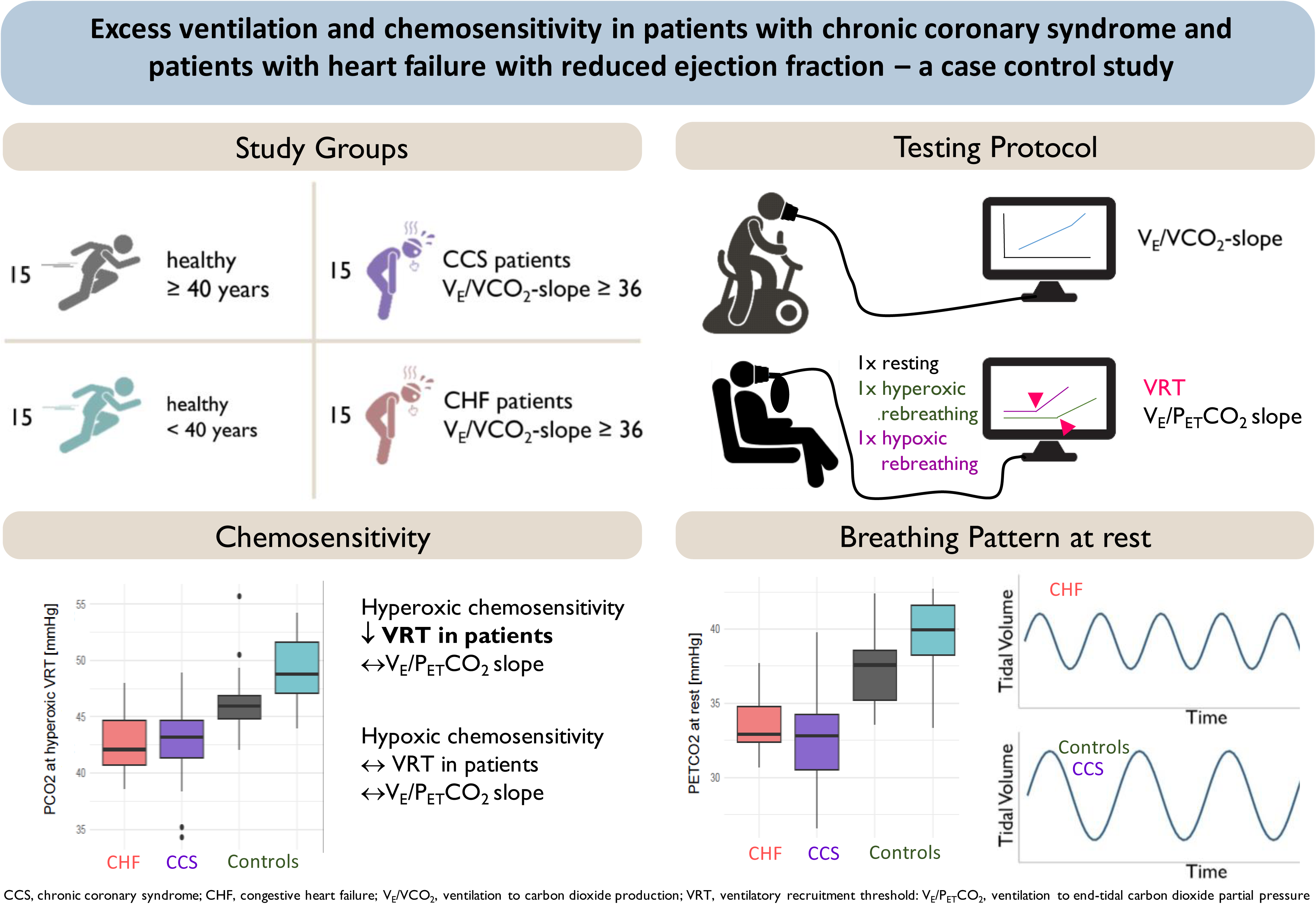

## Introduction

Chronic coronary syndrome (CCS) is the leading cause of heart failure (HF).^3^ An exaggerated ventilatory response to exercise, often accompanied by early exertional dyspnea, is a hallmark in patients with chronic heart failure (HF).^4, 5^ These observations have also been reported in patients with chronic coronary syndromes (CCS) and left ventricular dysfunction.^6^ Excess ventilation, often also termed ventilatory inefficiency^1^ has not only been associated with reduced exercise capacity and a reduced quality of life but also with poorer prognosis in both patients with HF and CCS.^7–11^ It is quantified by an increased V̇_E_/V̇CO_2_-slope, arising from an excessive rise of minute ventilation (V̇_E_) with respect to carbon dioxide production (V̇CO_2_) in the absence of metabolic acidosis.^12^ Based on the modified alveolar equation an increased V̇_E_/V̇CO_2_-slope can be explained by two factors: A reduced arterial CO_2_ partial pressure (P_a_CO_2_) and/or a high fraction of the tidal volume (V_T_) that goes to dead space (V_D_) (i.e., the V_D_/V_T_-ratio).^13^

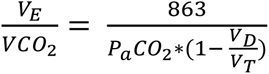

In patients with HF, impaired cardiac function may result in lung areas which are ventilated but poorly perfused (i.e., ventilation-perfusion mismatch) with V̇_E_ rising during exercise without sufficient rise in pulmonary perfusion.^14^ Furthermore, the V_D_/V_T_-ratio can be increased due to a reduced V_T_ during exercise when the diaphragm fatigues.^1^ Muscle fatigue in the diaphragm and/or the peripheral muscles leads to accumulating metabolites that trigger ergo reflexes which often elicit a steep increase in breathing frequency,^15^ resulting in a pattern of rapid shallow breathing (RSB).^14^ Increased chemosensitivity may further accelerate the abnormal ventilatory response to exercise in patients with HF ^4, 16^ and also in patients after acute myocardial infarction.^5^ Increased ventilatory drive during exercise has also been found in patients with left ventricular dysfunction but without established HF.^6^ Chemosensitivity, however, has not been assessed so far in patients with CCS.

Most early studies have assessed peripheral chemosensitivity to hypoxia by transient inhalation of pure nitrogen.^4, 16, 17^ However, while hypoxia may be present in some patients during sleep apnea,^18, 19^ hypoxia does not occur during exercise and is therefore unlikely to be related to inefficient ventilation. More recent studies have assessed respiratory chemosensitivity in patients with HF using isocapnic rebreathing to assess peripheral chemosensitivity to decreasing partial pressures of oxygen (PO_2_), and normoxic rebreathing to assess central chemosensitivity to increasing PCO_2_,^19, 20^ or Read’s rebreathing method.^3,5, 21^ Read’s rebreathing method has been refined by Duffin by introducing a 5 min hyperventilation period to lower arterial PCO_2_ so that the end tidal PCO_2_ at which V̇_E_ begins to rise (ventilatory recruitment threshold) can be identified additionally to chemosensitivity, which is the increase in ventilation with increasing CO_2_ during isoxia at hypoxic and hyperoxic tensions.^22–24^ This method has not been tested in patient populations so far but only in a single case study.^25^

The aims of the current study were to 1) compare central and peripheral respiratory chemoreflexes in terms of VRT and sensitivity measured by the Duffin rebreathing method in patients with inefficient ventilation and CCS to those with HF and to age-matched controls; 2) assess the contribution of central and peripheral chemoreflexes to ventilatory efficiency; 3) compare resting breathing patterns in these patient groups; 4) identify age-related differences in central and peripheral chemoreflexes by comparing healthy old to healthy young volunteers.

## Methods

### Study design and setting

This case-control study was conducted as a sub-study of the Breathe-HF trial (NCT05057884). Eligible patients with HF and CCS were identified and recruited during routine check-up visits at a tertiary university cardiovascular referral centre between April 2022 to April 2023. Healthy young and old volunteers were recruited by word of mouth. If they met the inclusion criteria and consented in writing, they were included in the study and measurements were performed as summarized in Supplement Figure 1. No follow-up was performed. The study was approved by the ethics committee of the Canton of Berne.

### Study participants

The study included four different groups of participants, two cardiac patient groups and two healthy control groups. Inclusion criteria for all groups were: age 18-80 years, capability of performing a cardiopulmonary exercise test on a cycling ergometer, willingness to participate in a study of a total 2 h duration, and provision of written informed consent. Exclusion criteria for all groups were: Present smoking, non-cardiac conditions and comorbidities associated with hyperventilation like pulmonary diseases, and pregnancy or lactation. Additional specific inclusion criteria for both patient groups were: Exertional dyspnea and V̇_E_/V̇CO_2_ slope≥36. A specific inclusion criterion for the CCS group was: no acute coronary syndrome in the last 3 months. Specific inclusion criteria for the HF group were: Left ventricular ejection fraction <50%, and optimal guideline-directed medical therapy for >3 months. A specific exclusion criterion for patients with CCS was: Diagnosis of heart failure. A specific exclusion criterion for patients with HF was: decompensation within the preceding 3 months. Exclusion criteria for healthy controls were: Past and present smoking, present consumption of aspirine, statins, beta blockers, alpha blocker, blockers of the renine-angiotensine-aldosterone system, Calcium-channel inhibitors, nitrates, nicorandil, ranolazine, phosphodiesterase-5 inhibitor, ivabradine, vitamin K antagonists, novel oral anticoagulants, glucocorticoids, beta mimetics.

Participants of the patient and the old control groups were recruited successively for groups to be comparable with regard to age and sex. The young control group was added for comparative purposes only and to assess the effect of age.

### Study Procedures

#### Body Composition Measurement

Body composition was assessed by bioelectrical impedance with a body composition analyzer (inBody 720, best4health gmbh, Bassersdorf, Switzerland). Weight, lean muscle mass, and body fat percentage were measured. Moreover, body mass index (BMI) was calculated.

#### Chemosensitivity Measurement

Central and peripheral chemosensitivity was assessed using the rebreathing protocol according to Duffin.^22^ This protocol was chosen because it can directly and reliably determine the ventilatory recruitment threshold (VRT), namely the P_ET_CO_2_ during hyper- and hypoxic conditions above which ventilation starts to increase, by establishing a PCO_2_ equilibrium between the alveolar air and the rebreathing bag. In contrast, other methods extrapolate the regression line of the ventilation to P_ET_CO_2_ slope to the abscissa, which creates larger errors.^26^ The advantages of this method compared to a conventional transient hypoxic test for the determination of peripheral chemosensitivity has been pointed out by Keir and colleagues.^27^

Patients were previously told to abstain from caffeine on the morning of the tests. The room was dimly lit and at a comfortable 22 degrees Celsius. The rebreathing procedure was performed using the Innocor system (COSMED Nordic ApS, Odense S, Denmark) running on a Windows XP embedded operating system on an integrated computer and a pulse oximeter (NONIN, sampling frequency 100 Hz) for O_2_ saturation. During measurements, participants wore an EU certified breathing mask (V2 Mask, Hans Rudolph, Shawnee, USA) as used during spiro-ergometries, covering nose and mouth. While most rebreathing procedures in literature used a mouthpiece and nose clip, Keir et al. demonstrated the feasibility of using a breathing mask for rebreathing tests.^28^ The breathing mask bears the advantage to allow the participants to breathe through nose or mouth as desired.

Once participants were fitted with the mask, they rested in a comfortable chair for 10 min. Breath-by-breath data was collected for the following parameters during resting and rebreathing: Oxygen consumption relative to body weight (V̇O_2_), CO_2_ production (V̇CO_2_), ventilation (V̇_E_), tidal volume (V_T_) and breathing frequency (BF). V̇_E_ and V_T_ were also adjusted to body surface are (BSA). End-tidal partial pressures of oxygen (P_ET_O_2_) and carbon dioxide (P_ET_CO_2_) were measured during 100 ms of highest O_2_ and CO_2_ values during each expiration. Instead of the proposed hyperventilation of 5 min by Duffin and colleagures,^22^ a duration of 2 min of hyperventilation was chosen, based on the findings by Boulet et al..^29^ After a 2 min hyperventilation and a decreased P_ET_CO_2_ by at least 10 mmHg below resting measurement, participants exhaled completely and a 3-way bi-directional valve (2100 Series, Hans Rudolph Inc.) was switched manually to connect patients with the rebreathing bag. Before starting the test, the 6-litre rebreathing bag was filled to three quarters of its volume with a gas mixture of 24% O_2_, 6% CO_2_ and balanced by N_2_. Participants were asked to take three deep breaths to reach an equilibrium between PCO_2_ in the rebreathing bag, lungs, arterial blood and mixed-venous blood.^22, 30^ After this, participants were instructed to breathe calmly and comfortably. By a manually controlled flow of 100% O_2_ to the rebreathing bag isoxia was kept at a P_ET_O_2_ of 150 mmHg. Rebreathing was terminated when P_ET_CO_2_ reached 60 mmHg or upon the participant’s request (by a previously agreed hand sign).^28^

The participant then rested for 15 min without the mask breathing room air.^31^ During this time, the bag was washed out three times and refilled to three quarters with a hypoxic gas mixture of 4.5% O_2_, 6% CO_2_ and balanced by N_2_. The hypoxic rebreathing was performed following the same protocol as before but P_ET_O_2_ was kept constant at 50 mmHg. The order of tests was kept constant for all participants, to avoid the effects of hypoxia on chemosensitivity that can last several hours.^32^ Parameters of the tenth minute of the resting period before the hyperoxic test (always first order) were averaged for resting values. For this period, the rapid shallow breathing index (RSBI) was also calculated as BF/V_T_ and BF/V_T_ relative to BSA.

Data analysis was performed as described by Duffin.^33^ Breath-by-breath P_ET_CO_2_ was plotted against time and fitted with a least-squares regression line. In order to minimize inter-breath variability, the equation for this line provided a predicted value of P_ET_CO_2_ against which V̇_E_ was plotted. By fitting a segmented linear regression model with a single breakpoint, the ventilatory recruitment threshold (VRT) after which V̇_E_ increased, and the V̇_E_/P_ET_CO_2_ slope starting at the VRT was determined (Duffin et al., 2000). A subtraction of the hypoxic test slope from the hyperoxic test slope in everyone was used to estimate the peripheral chemoreflex sensitivity.

#### Cardiopulmonary Exercise Testing

Exercise capacity was assessed with a CPET on a cycle ergometer. Prior to the test, a spirometry to determine forced vital capacity (FCV, l) and forced expiratory volume in one second (FEV_1_, l*min^-1^). Then, after sitting on the ergometer quietly for 3 min, blood pressure was measured two times and the lowest measurement was recorded. A 3 min warm-up was followed by an individually set ramp. Volumes, flows and gases were sampled continuously in an open spirometric system (Quark, Cosmed, Rome, Italy) and averaged over 8 breaths. Measured variables included V̇O_2_, carbon dioxide production (V̇CO_2_ ml*min^-1^), V̇_E_, BF, V_T_, and P_ET_O_2_ and P_ET_CO_2_, heart rate (HR, beats*min^-1^) and oxygen saturation (SpO_2_, %). V̇O_2peak_ (ml*min^-1^*kg^-1^) was defined as the highest value of oxygen consumption averaged over 30 s. The first (VT1) and second ventilatory threshold (VT2) were identified using the Wassermann’s 9-panel plot.^34^ The V̇_E_/V̇CO_2_-slope was determined from the start of the ramp until VT2. Further, the nadir of the V̇_E_/V̇CO_2_-ratio was defined as the lowest V̇_E_/V̇CO_2_-ratio during exercise.

#### Statistical Analysis

All analyses were performed by R (R Core Team, 2021, Version 4.1.0). CCS, HF, old and young controls were defined as exposures. Primary outcome was central and peripheral VRT and chemosensitivity. Secondary outcomes were rapid shallow breathing index (RSBI) and P_ET_CO_2_ at rest.

Baseline characteristics were tested between groups by Kruskal-Wallis tests followed by post-hoc testing adjusted for multiple testing by Benjamini-Hochberg correction. Categorical variables were tested by Fisher’s exact tests. Associations between variables were assessed by linear regression. Statistical significance for all tests was set at a p-value <0.05.

## Results

### Study Population

Each group included 15 participants (Supplement Figure 1). Of 53 patients with CCS qualifying for the study (32.8% of screened patients had V̇_E_/V̇CO_2_-slope ≥36), 20 could not be reached during the study period and 18 declined participation, leaving 15 who participated in the study. Of patients with HF, 59 qualified for inclusion (29.2% of screened patients had V̇_E_/V̇CO_2_-slope ≥36). Eighteen could not be reached during the study period and 26 declined participation in the study. Within the HF group, eight patients were classified as having reduced (HFrEF), and seven as having mildly reduced (HFmrEF).^35^ Fifteen old and young controls could be recruited. There were no significant differences between old healthy control subjects and the two patient groups with regard to baseline characteristics (Table 1). The only significantly different baseline characteristics were age and percent body fat between old and young healthy control subjects.

**Table 1:** Baseline characteristics of the two patient and two healthy groups.

|  | **CHF patients**<br>**(n = 15)** | **CCS patients**<br>**(n = 15)** | **Old controls**<br>**(n = 15)** | **Young controls**<br>**(n = 15)** |
| --- | --- | --- | --- | --- |
| Age | 68 (63, 71) | 68 (63, 73) | 68 (60, 72) | 25 (24, 30)* |
| Female n (%) | 4 (26.7) | 3 (20.0) | 4 (26.7) | 6 (40.0) |
| Weight [kg] | 79.9 (66.0, 84.7) | 76.4 (72.0, 83.6) | 76.6 (66.6, 84.3) | 68.2 (63.4, 73.9) |
| Height [cm] | 173 (164, 178) | 174 (166, 178) | 177 (166, 182) | 173 (169, 178) |
| BMI [kg/m<sup>2</sup>] | 26.2 (23.4, 28.8) | 27.9 (23.1, 29.2) | 25.2 (23.0, 26.1) | 22.2 (21.0, 23.6) |
| Muscle mass [kg]* | 36.0 (31.1, 40.5) | 31.9 (30.0, 33.9) | 32.1 (27.5, 36.9) | 32.5 (28.6, 35.8) |
| Percent body fat [%]<sup>a</sup> | 27.0 (22.1, 31.9) | 30.1 (23.5, 32.5) | 22.8 (19.7, 26.0) | 16.9 (13.2, 18.8)* |
| Systolic BP [mmHg] | 110 (100, 119) | 120 (115, 135) | 120 (113, 120) | 117 (110, 120) |
| Diastolic BP [mmHg] | 70 (63, 74)* | 79 (65, 80) | 80 (78, 83) | 80 (76, 80) |
| MIP [cmH<sub>2</sub>O]<sup>b</sup> | 83 (57, 98) | 69 (58, 89) |  |  |
| NYHA class I/II | 3/12 | 7/8 |  |  |
| Presence of EOv | 5 (33.3) | 2 (13.3) |  |  |
| LV ejection fraction [%] | 40 (36, 47) | 60 (57, 64) |  |  |
| LAVI [mL/m<sup>2</sup>] | 38.5 (31.4, 44.4) | 29.9 (25.4, 33.3) |  |  |
| LVEDVi [mL/m<sup>2</sup>] | 68.0 (54.5, 98.2) | 46.4 (45.1, 53.9) |  |  |
| History of ACS (%) | 8 (53.3) | 10 (66.7) |  |  |
| History of PCI (%) | 9 (60.0) | 11 (73.3) |  |  |
| Cardiac implantable electric device (%) | 9 (60.0) | 1 (6.7) |  |  |
| *Medication* |  |  |  |  |
| ACEi/AT2 (%) | 3 (20.0) | 13 (86.7) |  |  |
| ARNI (%) | 9 (60.0) | 0 (0.0) |  |  |
| MRA (%) | 8 (53.3) | 0 (0.0) |  |  |
| SGLT2i (%) | 14 (93.3) | 4 (26.7) |  |  |
| Betablocker (%) | 12 (80.0) | 12 (80.0) |  |  |
| Carvedilol (%) | 0 (0) | 0 (0) |  |  |

Data are indicated as median (1<sup>st</sup>, 3<sup>rd</sup> quartiles) or number of subjects (%).
\* Adjusted p-value <0.05 of post hoc Kruskal Wallis tests between old control subjects and other groups
<sup>a</sup> Data missing from one CCS and six CHF patients due to inability to conduct body composition measurement because of Cardiac implantable electric device (CIED).
<sup>b</sup> Data available only from 9 CHF and 11 CCS patients.
CHF, chronic heart failure; CCS, acute/chronic coronary syndrome; BMI, body mass index; BP, blood pressure; MIP, maximal inspiratory pressure; NYHA class, New York Heart Association class; EOv, exercise oscillatory ventilation; LV, left ventricular; LAVI, left atrial volume index; LVEDVi, left ventricle end-diastolic volume index; ACS, acute coronary syndrome; PCI, Percutaneous coronary intervention; ACEi, Angiotensin-converting-enzyme inhibitors; AT2, angiotensin II type 2 receptor agonist; ARNI, angiotensin receptor-neprilysin inhibitor; MRA, mineralocorticoid receptor antagonist; SGLT2i, mineralocorticoid receptor antagonist inhibitor

### Results of Chemosensitivity Tests

There was mask leakage in one old healthy control during the hyperoxic test and in another old healthy control in the hypoxic test, leaving results of only 14 subjects in this group for data analysis. During the hypoxic test, there was mask leakage in two patients with HF and four patients with CCS. Further, one patient with HF and two patients with CCS stopped the hypoxic tests after 3-5 breaths, which did not allow the determination of VRT and slope. Therefore, only 12 patients were included for data analysis of the hypoxic test. A typical example of the sampled data of the two tests in one patient with HF and one age- and sex-matched control is shown in Supplement Figure 2.

The HF and CCS patient groups had significantly reduced median hyperoxic test VRTs compared to old controls (p=0.004 and 0.01, respectively, Figure 1a, Table 2), but hypoxic test VRTs did not differ (Figure 1b, Table 2). Young healthy controls had higher median VRTs than old controls for both hyper- and hypoxic tests, which were not quite significant (p=0.06 and p=0.07, respectively).

**Figure 1:**
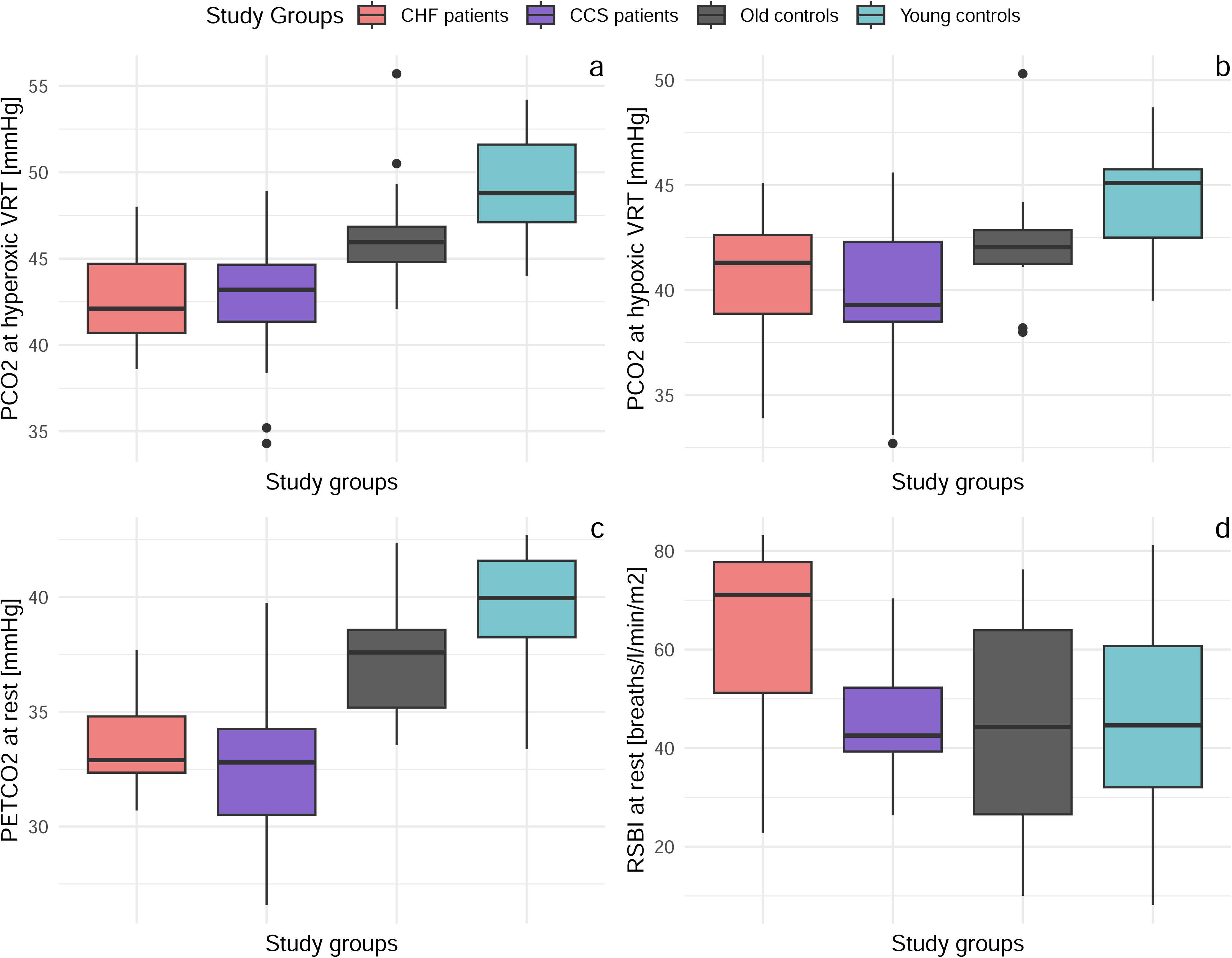
Boxplots of the four study groups showing P_ET_CO_2_ at hyperoxic VRT (a), P_ET_CO_2_ at hypoxic VRT (b), P_ET_CO_2_ at rest (c), and RSBI (d). VRTs were determined from rebreathing and RSBI and P_ET_CO_2_ from resting measurement before rebreathing. CHF, chronic heart failure; CCS, chronic coronary syndrome; PCO_2_, endtidal carbon dioxide pressure; VRT, ventilatory response threshold; RSBI, rapid shallow breathing index; P_ET_CO_2_, endtidal carbon dioxide pressure

**Table 2:**
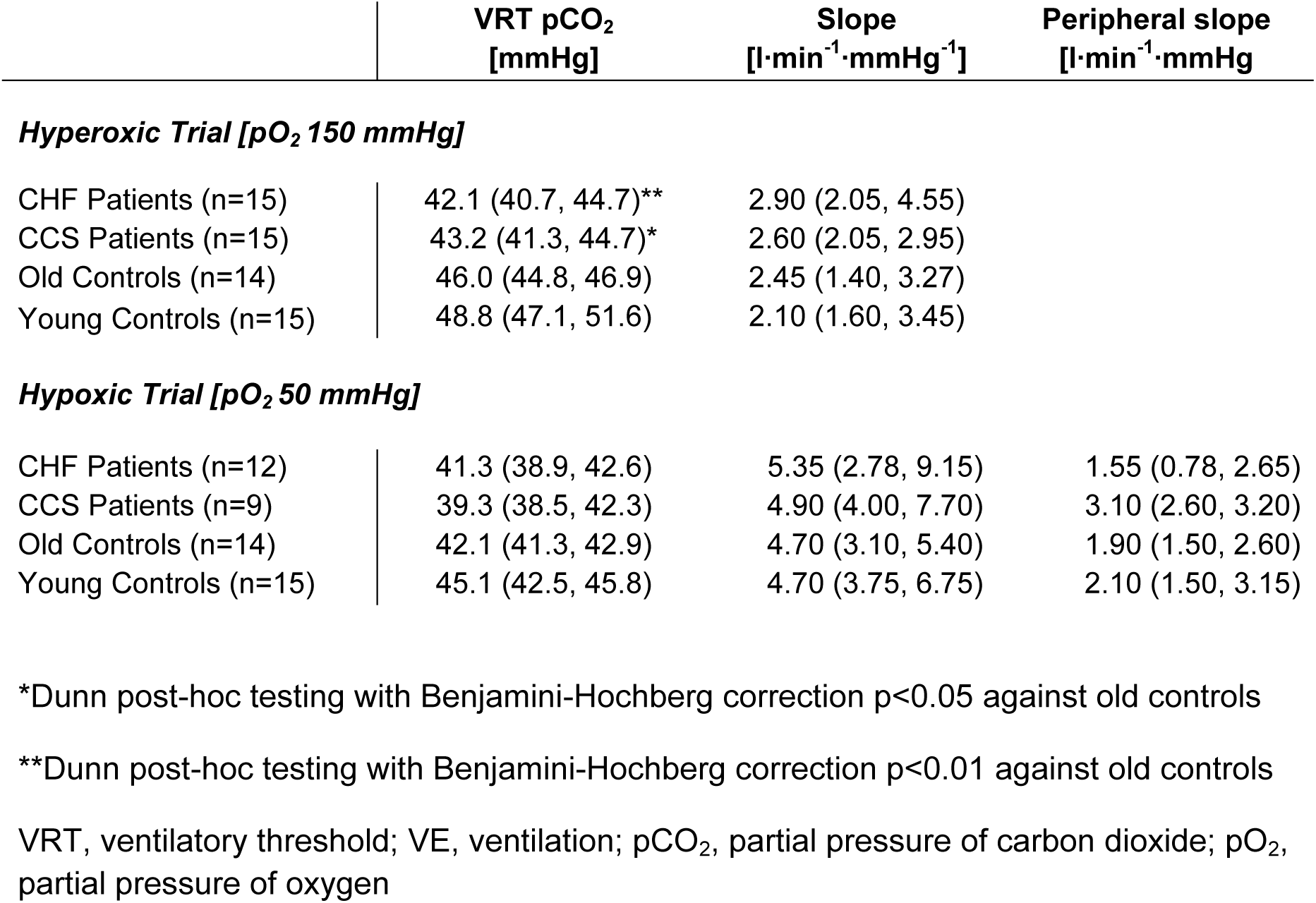
Parameters measured with hyperoxic and hypoxic rebreathing.

|  | VRT pCO<sub>2</sub><br>[mmHg] | Slope<br>[l·min<sup>-1</sup>·mmHg<sup>-1</sup>] | Peripheral slope<br>[l·min<sup>-1</sup>·mmHg] |
| --- | --- | --- | --- |
| ***Hyperoxic Trial [pO<sub>2</sub> 150 mmHg]*** |  |  |  |
| CHF Patients (n=15) | 42.1 (40.7, 44.7)** | 2.90 (2.05, 4.55) |  |
| CCS Patients (n=15) | 43.2 (41.3, 44.7)* | 2.60 (2.05, 2.95) |  |
| Old Controls (n=14) | 46.0 (44.8, 46.9) | 2.45 (1.40, 3.27) |  |
| Young Controls (n=15) | 48.8 (47.1, 51.6) | 2.10 (1.60, 3.45) |  |
| ***Hypoxic Trial [pO<sub>2</sub> 50 mmHg]*** |  |  |  |
| CHF Patients (n=12) | 41.3 (38.9, 42.6) | 5.35 (2.78, 9.15) | 1.55 (0.78, 2.65) |
| CCS Patients (n=9) | 39.3 (38.5, 42.3) | 4.90 (4.00, 7.70) | 3.10 (2.60, 3.20) |
| Old Controls (n=14) | 42.1 (41.3, 42.9) | 4.70 (3.10, 5.40) | 1.90 (1.50, 2.60) |
| Young Controls (n=15) | 45.1 (42.5, 45.8) | 4.70 (3.75, 6.75) | 2.10 (1.50, 3.15) |

\*Dunn post-hoc testing with Benjamini-Hochberg correction p<0.05 against old controls
\*\*Dunn post-hoc testing with Benjamini-Hochberg correction p<0.01 against old controls
VRT, ventilatory threshold; VE, ventilation; pCO<sub>2</sub>, partial pressure of carbon dioxide; pO<sub>2</sub>, partial pressure of oxygen

In all groups, hypoxic test slopes were significantly higher than hyperoxic test slopes (all p<0.004, Table 2). However, hyperoxic and hypoxic test slopes (sensitivity, V̇_E_ vs. P_ET_CO_2_) were not significantly different between the patient groups and old controls (Table 2). Hyperoxic and hypoxic slopes were not different between young and old controls.

Linear relationships between V̇_E_/V̇CO_2_-slopes and hyperoxic as well as hypoxic VRT were only significant for the pooled sample (r^2^=0.35 and r^2^=0.24, both p<0.0001) but not within groups (Figure 2a and 2b). There were no linear relationships between V̇_E_/V̇CO_2_-slopes and hyperoxic and hypoxic test slopes of the rebreathing tests, within groups or in the pooled sample. However, there was a significant linear association between V̇_E_/V̇CO_2_-slopes and the maximal P_ET_CO_2_ reached during the CPET. This finding was true for both within groups (all p<0.02) as well as for the pooled sample (r^2^=0.79, p<0.0001, Figure 2c).

**Figure 2:**
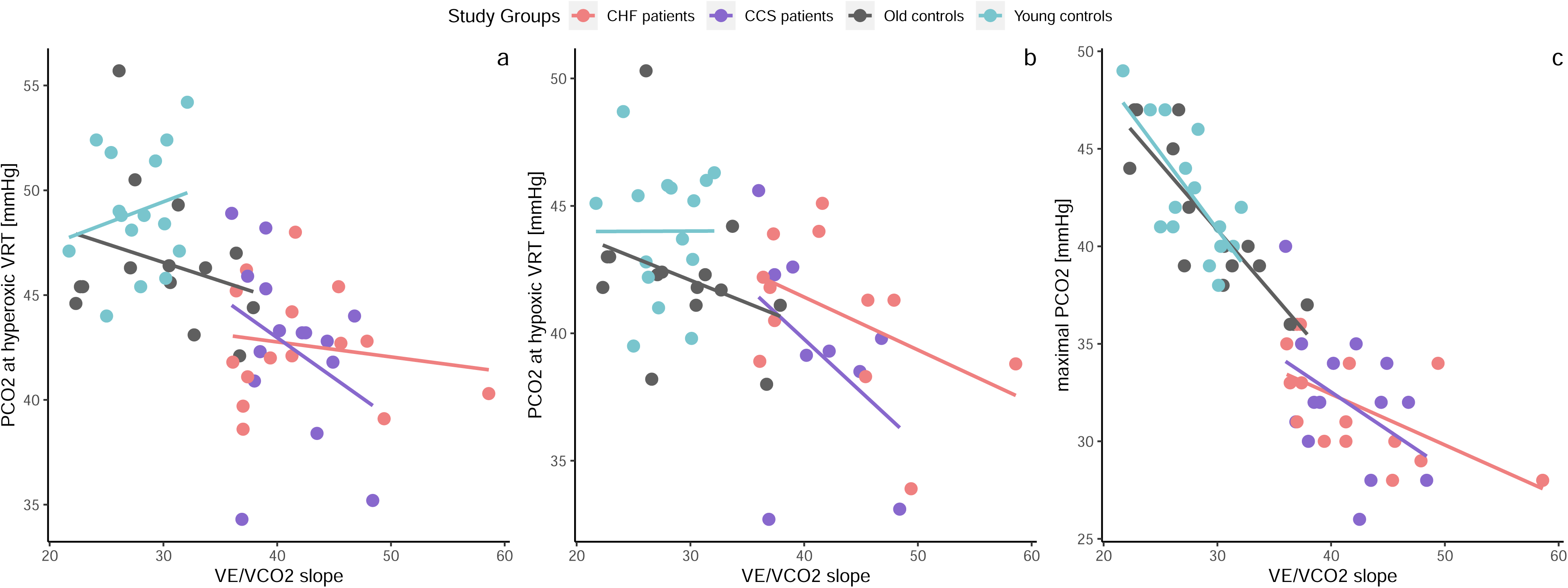
Within group linear regressions between V̇_E_/V̇CO_2_ slope of CPET and PCO_2_ at hyper- (a) and hypoxic (b) VRTs of the chemosensitivity measurements and maximal PCO_2_ of CPET (c). CHF, chronic heart failure; CCS, chronic coronary syndrome; V̇_E_, ventilation; V̇CO_2_, carbon dioxide production; PCO_2_, endtidal carbon dioxide pressure; VRT, ventilatory response threshold; CPET, cardiopulmonary exercise test

There was a significant linear correlation between hyperoxic and hypoxic VRT in the pooled sample (r^2^=0.68, p<0.0001) as well as within each group (all p≤0.05, Supplement Figure 3a). Correlations between hyper- and hypoxic slopes were significant for the pooled sample (r^2^=0.24, p<0.0001) and within all groups (p<0.006) except the CCS group (p=0.201, Supplement Figure 3b). P_ET_CO_2_ at rest was related to hyperoxic and hypoxic test VRTs within all groups except for the hyperoxic VRT of the young healthy group (Figures 3a and 3b).

**Figure 3:**
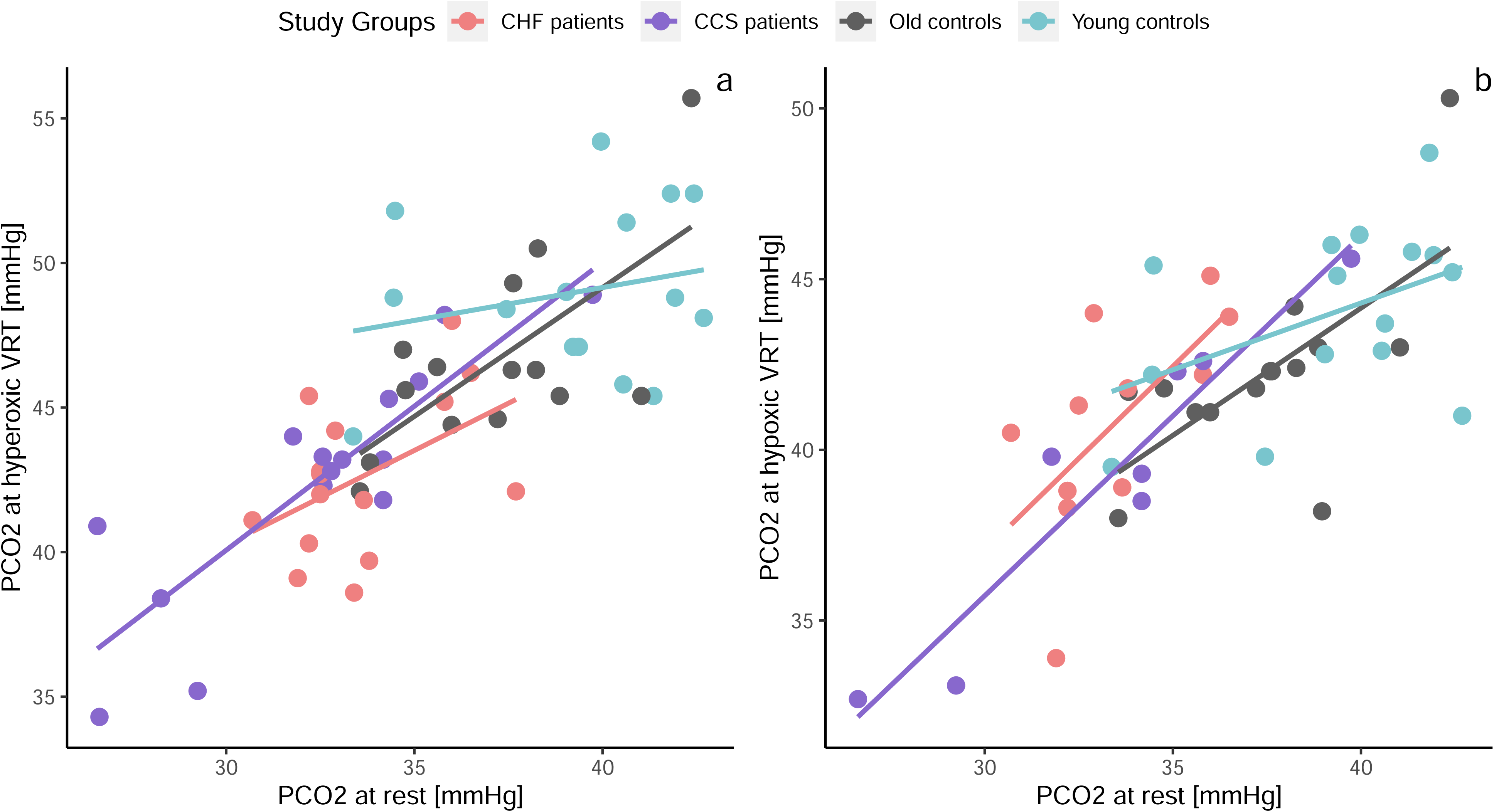
Within group linear regressions between resting P_ET_CO_2_ and hyper- (a) as well as hypoxic VRT (b). PCO_2_, endtidal carbon dioxide pressure; VRT, ventilatory response threshold

### Results of Resting Measurements and Cardiopulmonary Exercise Tests

Resting parameters of the two patient groups were comparable to old control subjects except for P_ET_CO_2_, which was lower in both patient groups (Figure 1c), and BF and RSBI (Figure 1d), which were higher in the HF group compared to old controls and to patients with CCS (Supplement Table 1). Based on the inclusion criteria for both patient groups, they had significantly higher V̇_E_/V̇CO_2_-slopes, nadir V̇_E_/V̇CO_2_ and lower maximal P_ET_CO_2_ (Supplement Table 1). At peak exercise, power, VO_2,_ absolute and relative V̇_E_, P_ET_CO_2_ and HR were lower than in the old control subjects (Supplement Table 1). Patients with HF had higher BF and RSBI than patients with CCS (p<0.05 for BF and RSBI relative to BSA, p=0.05 for RSBI) than patients with CCS.

## Discussion

### General

The present study selected patients with chronic HFrEF/HFmrEF or CCS who presented with exercise excess ventilation and compared their central and peripheral chemosensitivity and resting breathing pattern to age-matched and young healthy controls. This is the first study who included two cohorts of well phenotyped patients with HF (with reduced ejection fraction) and CCS, and who simultaneously assessed breathing efficiency at rest with a standardised tests as well as at exercise with CPET based measurements. Further, the inclusion of both sexes as well as a young healthy control group allowed a direct comparison to healthy subjects without ventilatory inefficiency as well as the effect of age on the measured parameters.

Key findings of the study were: 1) Patients with CCS and HF had lower hyperoxic rebreathing test VRTs compared to old and young controls. 2) Central and peripheral chemosensitivity quantified by V̇_E_/P_ET_CO_2_ slopes were not different between groups. 3) Both patient groups had also excess ventilation at rest with lower P_ET_CO_2_ compared to the control groups, which correlated with the decreased hyperoxic and hypoxic rebreathing test VRTs. 4) Patients with HF had a rapid shallow breathing pattern and higher BF and RSBI at rest compared to patients with CCS and healthy controls. This is the first study to show that excess ventilation, which has been well investigated with exercise and whose negative correlation with poor outcome has been well investigated in patients with HF, is also present at rest and also in patients with CCS. This suggests that excess ventilation in these patients may be at least in part be caused by factors other than congestion of the lung due to reduced cardiac function and not only be elicited by exercise.

### Chemoreflex Ventilatory Recruitment Threshold

A lower VRT in patients with HF has previously been found in patients with univentricular heart using both transient hypoxia and Read rebreathing tests.^36^ Reduced VRT reflects excess ventilation (e.g. altitude hypoxia^37^), which in this case likely results from high or normal anion gap metabolic acidosis and reduced bicarbonate.^38, 39^ Indeed, electrolyte derangement such as by hyponatriemia, hypocalcemia, hypokalemia, hypomagnesemia and hypophosphatemia, are common in patients with HF as documented by several studies.^40–42^ Importantly, central chemosensitivity has been found to be significantly reduced in patients with HF after intravenous iron infusion.^2^ In line with our study, they found no effect of ironinfusion on peripheral chemosensitivity. The fact that patients with CCS and HF had comparable values of reduced ventilatory efficiency, reduced VRT and reduced P_ET_CO_2_ at rest suggests that they had the same degree of excess ventilation although with a different breathing pattern in that only patients with HF developed a rapid shallow breathing pattern. However, in this study hyperoxic and hypoxic rebreathing test VRTs correlated poorly with ventilatory drive during exercise (V̇_E_/V̇CO_2_ slope), with the correlation significant only in the pooled group and not within individual groups.

Furthermore, hyperoxic and hypoxic rebreathing test V̇_E_/P_ET_CO_2_ slopes did not correlate with V̇_E_/V̇CO_2_ slopes. This finding is in contrast to a study by Tomita and colleagues who found a significant correlation between V̇_E_/V̇CO_2_ slope and V̇_E_/P_ET_CO_2_ slopes during hyperoxic Read rebreathing.^5^ Their hyperoxic testing procedure differed from ours in that PO_2_ decreased during the test while it was kept isoxic in ours. There are also other studies that found some association between V̇_E_/V̇CO_2_ slope and peripheral chemosensitivity^4^ and central chemosensitivity.^4, 19^ The weak contribution of chemosensitivity to exercise ventilatory efficiency suggest that other factors are important. A likely culprit for an elevated V̇_E_/V̇CO_2_ slope in patients with HF or CCS may be the ergoreflex.^43^ Scott and colleagues found a similar relationship (r=0.576, p<0.01) between V̇_E_/V̇CO_2_ slope and increase in ventilation above rest at 2 min after hand grip exercise and post-exercise regional circulatory occlusion, a test quantifying metaboreflex, in 15 patients with CHF and 8 healthy controls.^44^ Our results indicate that the direct contribution of chemosensitivity to exercise ventilatory efficiency is minimal in both heart patients and healthy controls but that V̇_E_/V̇CO_2_ slope is related to maximally tolerated P_ET_CO_2_.

The age related decrease in VRT for both hyperoxic and hypoxic rebreathing tests observed in our healthy groups corresponds to findings of earlier studies.^45^ Garcia-Rio and colleagues found the VRTs of both, hyperoxic hypercapnic and isocapnic hypoxic stimulation in elderly subjects to be at a lower P_ET_CO_2_ than in young healthy subjects.^45^ However, they found that the hyperoxic hypercapnic VRT increased again after age 75.

### Chemoreflex sensitivity

Our findings of comparable VE/P_ET_CO_2_ slopes of the hyper- and hypoxic rebreathing between our patient and healthy groups are in contrast to some previous studies who have reported increased central chemosensitivity to hypercapnia^19, 20, 46, 47^ and peripheral chemosensitivity to hypoxia in heart failure,^16, 19, 20, 46, 48^ Tomita and colleagues^5^ found increased chemosensitivity in patients with acute myocardial infarction using Read hyperoxic rebreathing tests, and peripheral chemosensitivity measured using the Duffin isoxic rebreathing method has been found to be associated with cardiovascular risk in a Chinese population.^49^

We found no differences between hyperoxic and hypoxic slopes (sensitivity) between young and old controls in agreement with previous findings. Increased chemosensitivity with increasing age was found during sleep,^50^ but no differences between healthy younger and older men were found with transient hypoxia by rebreathing of pure nitrogen by another study.^51^

### Resting P_ET_CO2

While excess ventilation with exercise has been well studied, resting excess ventilation has rarely been investigated. Resting P_a_CO_2_ <31 mmHg has been found to be associated with increased all-cause mortality.^52^ We have recently shown that increased resting breathing frequency has been found to be associated with major adverse events in patients with left ventricular dysfunction.^6^ The association of a lower resting P_a_CO_2_ with VRT or chemosensitivity has not been investigated, although in patients with HF it has been found that Cheyenne Stokes respiration was associated with a lower resting P_a_CO_2_.^53^ It has been suggested that low P_a_CO_2_ in patients with HF may be a respiratory manifestation of elevated left ventricular filling pressures.^54^ However, since we found the same relationships between VRT and resting P_ET_CO_2_ in all groups (with healthy groups and patients with CCS not having elevated left ventricular filling pressures), our data suggests that there may be a neurological or hormonal rather than a circulatory cause for the resting excess ventilation in patients with HF and CCS.

### Limitations

As in a recent validation study,^26^ we found the Duffin hyperoxic and hypoxic rebreathing tests to be feasible in healthy controls, but less feasible in patients with CCS or HF. One problem particularly during the hypoxic test was that after hyperventilation and the subsequent three breaths for equilibration, many patients took a break from breathing for a few seconds to recover from hyperventilation. When breathing resumed they had already surpassed their VRT and stopped the test after only a few more breaths. The dearth of breaths reduced the quality of VRT and V̇_E_/P_ET_CO_2_ slope determinations. Mask leakage was an additional problem in our study, however, sealing the lips around a mouthpiece can also be challenging. The consequent exclusion of some hypoxic rebreathing tests may have led to underpowering of the hypoxic test. Further, since we only included patients with V̇_E_/V̇CO_2_ slopes ≥36, we cannot extrapolate our findings of similarly altered chemoreflexes and reduced P_ET_CO_2_ in patients with CCS and HF to patients with only mildly increased ventilatory inefficiency. Last but not least, we only measured the ventilatory response to increasing values of P_ET_CO_2_. Due to the absence of simultaneous blood gas analysis, we assumed that P_ET_CO_2_ reflected arterial PCO_2_, an assumption supported by recent experimental evidence.^55^

## Conclusions

The present study shows that patients with HF or CCS and excess ventilation during exercise also have excess ventilation at rest. Further, we show that patients with CCS have similarly reduced VRT during hyperoxic rebreathing as patients with HFrEF/HFmrEF. The increased ventilatory drive during exercise may be related more to VRT than to chemosensitivity and hence be a symptom of electrolyte derangement. An increased ventilatory drive produced by a lower VRT likely reduces the maximal P_ET_CO_2_ that can be tolerated during exercise. These associations were found in both patient groups, although the pattern of excess ventilation was different in patients with HF in that they adopted by rapid shallow breathing while patients with CCS showed a normal breathing pattern.

## Data Availability

Original coded data will be made available upon direct request to the corresponding author.

## Conflict of interest statement

None of the authors have any conflict of interest to declare.

## Funding information

No funding was received for this study.

## Non-standardised abbreviations

BMI: body mass index
BF: breathing frequency
CCS: chronic coronary syndrome
CO_2_: carbon dioxide
CPET: cardiopulmonary exercise test
HF: heart failure
HFpEF: heart failure with preserved ejection fraction
HFmrEF: heart failure with mildly reduced ejection fraction
HFrEF: heart failure with reduced ejection fraction
HR: heart rate
O_2_: oxygen
P_a_CO_2_: arterial partial pressure of carbon dioxide
P_a_O_2_: arterial partial pressure of oxygen
P_ET_CO_2_: end-tidal partial pressure of carbon dioxide
P_ET_O_2_: end-tidal partial pressure of oxygen
RER: V̇CO_2_/V̇O_2_
RPE: rate of perceived exertion
RSBI: rapid shallow breathing index
SpO_2_: oxygen saturation
V_D_: pulmonary dead space
V̇_E_: ventilation
V̇CO_2_: carbon dioxide production
V̇O_2_: oxygen production
V_T_: tidal volume
VT1: first ventilatory threshold
VT2: second ventilatory threshold
VRT: ventilatory recruitment threshold

---

Supplement Figure 1: Patient/participant flow of the four groups. CHF, chronic heart failure; CCS, chronic coronary syndrome

Supplement Figure 2: Breath-by-breath data of a typical male patient with HF (left) and male healthy age-matched control (right). Shown are the VE at increasing P_ET_CO_2_ of the hyper-(green symbols) and hypoxic (blue symbols) rebreathing tests in the same plot. Additionally, P_ET_CO_2_ P_ET_CO_2_ and S_a_O_2_ is shown of the hypoxic test, as S_a_O_2_ only decreases in the hypoxic but not the hyperoxic rebreathing test. HF, heart failure; P_ET_CO_2,_ endtidal carbon dioxide partial pressure; S_a_O_2_, oxygen saturation; VE, ventilation

Supplement Figure 3: Within group linear regressions between PCO_2_ at hyper- and hypoxic VRTs (a) and hyper- and hypoxic V̇_E_/PCO_2_ slopes (b) of the chemosensitivity measurements. CHF, chronic heart failure; CCS, chronic coronary syndrome; V̇_E_, ventilation; PCO_2_, endtidal carbon dioxide pressure; VRT, ventilatory response threshold

## Notes

### Competing Interest Statement

The authors have declared no competing interest.

### Clinical Trial

NCT05057884

### Funding Statement

There was no funding for the present study.

### Author Declarations

Ethikkommission des Kt. Bern

## References

1. Neder JA, Phillips DB, O’Donnell DE, et al. Excess ventilation and exertional dyspnoea in heart failure and pulmonary hypertension. The European respiratory journal 2022; 60(5).

2. Caravita S, Faini A, Vignati C, et al. Intravenous iron therapy improves the hypercapnic ventilatory response and sleep disordered breathing in chronic heart failure. European journal of heart failure 2022; 24(10): 1940–9.

3. Ziaeian B, Fonarow GC. Epidemiology and aetiology of heart failure. Nat Rev Cardiol 2016; 13(6): 368–78.

4. Chua TP, Clark AL, Amadi AA, et al. Relation between chemosensitivity and the ventilatory response to exercise in chronic heart failure. Journal of the American College of Cardiology 1996; 27(3): 650–7.

5. Tomita T, Takaki H, Hara Y, et al. Attenuation of hypercapnic carbon dioxide chemosensitivity after postinfarction exercise training: possible contribution to the improvement in exercise hyperventilation. Heart 2003; 89(4): 404–10.

6. Eser P, Marcin T, Prescott E, et al. Breathing pattern and pulmonary gas exchange in elderly patients with and without left ventricular dysfunction-modification with exercise-based cardiac rehabilitation and prognostic value. Front Cardiovasc Med 2023; 10: 1219589.

7. Ponikowski PP, Chua TP, Francis DP, et al. Muscle ergoreceptor overactivity reflects deterioration in clinical status and cardiorespiratory reflex control in chronic heart failure. Circulation 2001; 104(19): 2324–30.

8. Arena R, Myers J, Aslam SS, et al. Peak VO2 and VE/VCO2 slope in patients with heart failure: a prognostic comparison. American heart journal 2004; 147(2): 354–60.

9. Nadruz W, Jr., West E, Sengelov M, et al. Prognostic Value of Cardiopulmonary Exercise Testing in Heart Failure With Reduced, Midrange, and Preserved Ejection Fraction. Journal of the American Heart Association 2017; 6(11).

10. Chua TP, Ponikowski P, Harrington D, et al. Clinical correlates and prognostic significance of the ventilatory response to exercise in chronic heart failure. Journal of the American College of Cardiology 1997; 29(7): 1585–90.

11. Nayor M, Xanthakis V, Tanguay M, et al. Clinical and Hemodynamic Associations and Prognostic Implications of Ventilatory Efficiency in Patients With Preserved Left Ventricular Systolic Function. Circulation Heart failure 2020; 13(5): e006729.

12. Agostoni P, Guazzi M. Exercise ventilatory inefficiency in heart failure: some fresh news into the roadmap of heart failure with preserved ejection fraction phenotyping. European journal of heart failure 2017; 19(12): 1686–9.

13. Wang MC, Corbridge TC, McCrimmon DR, et al. Teaching an intuitive derivation of the clinical alveolar equations: mass balance as a fundamental physiological principle. Adv Physiol Educ 2020; 44(2): 145–52.

14. Weatherald J, Sattler C, Garcia G, et al. Ventilatory response to exercise in cardiopulmonary disease: the role of chemosensitivity and dead space. The European respiratory journal 2018; 51(2).

15. Piepoli M, Clark AL, Volterrani M, et al. Contribution of muscle afferents to the hemodynamic, autonomic, and ventilatory responses to exercise in patients with chronic heart failure: effects of physical training. Circulation 1996; 93(5): 940–52.

16. Ponikowski P, Chua TP, Piepoli M, et al. Augmented peripheral chemosensitivity as a potential input to baroreflex impairment and autonomic imbalance in chronic heart failure. Circulation 1997; 96(8): 2586–94.

17. Ponikowski P, Chua TP, Anker SD, et al. Peripheral chemoreceptor hypersensitivity: an ominous sign in patients with chronic heart failure. Circulation 2001; 104(5): 544–9.

18. Giannoni A, Gentile F, Navari A, et al. Contribution of the Lung to the Genesis of Cheyne-Stokes Respiration in Heart Failure: Plant Gain Beyond Chemoreflex Gain and Circulation Time. Journal of the American Heart Association 2019; 8(13): e012419.

19. Giannoni A, Emdin M, Poletti R, et al. Clinical significance of chemosensitivity in chronic heart failure: influence on neurohormonal derangement, Cheyne-Stokes respiration and arrhythmias. Clinical science (London, England : 1979) 2008; 114(7): 489-97.

20. Giannoni A, Emdin M, Bramanti F, et al. Combined increased chemosensitivity to hypoxia and hypercapnia as a prognosticator in heart failure. Journal of the American College of Cardiology 2009; 53(21): 1975–80.

21. Read DJ. A clinical method for assessing the ventilatory response to carbon dioxide. Australas Ann Med 1967; 16(1): 20–32.

22. Duffin J. Measuring the respiratory chemoreflexes in humans. Respir Physiol Neurobiol 2011; 177(2): 71–9.

23. Duffin J. Measuring the ventilatory response to hypoxia. The Journal of physiology 2007; 584(Pt 1): 285–93.

24. Mohan RM, Amara CE, Cunningham DA, et al. Measuring central-chemoreflex sensitivity in man: rebreathing and steady-state methods compared. Respir Physiol 1999; 115(1): 23–33.

25. Keir DA, Duffin J, Badrov MB, et al. Hypercapnia During Wakefulness Attenuates Ventricular Ectopy: Observations in a Young Man With Heart Failure With Reduced Ejection Fraction. Circulation Heart failure 2020; 13(6): e006837.

26. Jensen D, Mask G, Tschakovsky ME. Variability of the ventilatory response to Duffin’s modified hyperoxic and hypoxic rebreathing procedure in healthy awake humans. Respir Physiol Neurobiol 2010; 170(2): 185–97.

27. Keir DA, Duffin J, Floras JS. Measuring Peripheral Chemoreflex Hypersensitivity in Heart Failure. Frontiers in physiology 2020; 11: 595486.

28. Keir DA, Duffin J, Millar PJ, et al. Simultaneous assessment of central and peripheral chemoreflex regulation of muscle sympathetic nerve activity and ventilation in healthy young men. The Journal of physiology 2019; 597(13): 3281–96.

29. Boulet LM, Tymko MM, Jamieson AN, et al. Influence of prior hyperventilation duration on respiratory chemosensitivity and cerebrovascular reactivity during modified hyperoxic rebreathing. Exp Physiol 2016; 101(7): 821–35.

30. Jensen D, Wolfe LA, O’Donnell DE, et al. Chemoreflex control of breathing during wakefulness in healthy men and women. J Appl Physiol (1985) 2005; 98(3): 822-8.

31. Duffin J, McAvoy GV. The peripheral-chemoreceptor threshold to carbon dioxide in man. The Journal of physiology 1988; 406: 15–26.

32. Mateika JH, Mendello C, Obeid D, et al. Peripheral chemoreflex responsiveness is increased at elevated levels of carbon dioxide after episodic hypoxia in awake humans. J Appl Physiol (1985) 2004; 96(3): 1197-205; discussion 6.

33. Duffin J, Mohan RM, Vasiliou P, et al. A model of the chemoreflex control of breathing in humans: model parameters measurement. Respir Physiol 2000; 120(1): 13–26.

34. Marcin T, Eser P, Prescott E, et al. Training intensity and improvements in exercise capacity in elderly patients undergoing European cardiac rehabilitation - the EU-CaRE multicenter cohort study. PloS one 2020; 15(11): e0242503.

35. McDonagh TA, Metra M, Adamo M, et al. 2021 ESC Guidelines for the diagnosis and treatment of acute and chronic heart failure. European heart journal 2021; 42(36): 3599-726.

36. Chua TP, Iserin L, Somerville J, et al. Effects of chronic hypoxemia on chemosensitivity in patients with univentricular heart. Journal of the American College of Cardiology 1997; 30(7): 1827–34.

37. Frost S, Pham K, Puvvula N, et al. Changes in hypoxic and hypercapnic ventilatory responses at high altitude measured using rebreathing methods. J Appl Physiol (1985) 2024.

38. Fenves AZ, Emmett M. Approach to Patients With High Anion Gap Metabolic Acidosis: Core Curriculum 2021. Am J Kidney Dis 2021; 78(4): 590–600.

39. Ren X, Robbins PA. Ventilatory responses to hypercapnia and hypoxia after 6 h passive hyperventilation in humans. The Journal of physiology 1999; 514 (Pt 3)(Pt 3): 885-94.

40. Milionis HJ, Alexandrides GE, Liberopoulos EN, et al. Hypomagnesemia and concurrent acid-base and electrolyte abnormalities in patients with congestive heart failure. European journal of heart failure 2002; 4(2): 167–73.

41. Urso C, Brucculeri S, Caimi G. Acid-base and electrolyte abnormalities in heart failure: pathophysiology and implications. Heart failure reviews 2015; 20(4): 493–503.

42. Dei Cas L, Metra M, Leier CV. Electrolyte disturbances in chronic heart failure: metabolic and clinical aspects. Clinical cardiology 1995; 18(7): 370–6.

43. Aimo A, Saccaro LF, Borrelli C, et al. The ergoreflex: how the skeletal muscle modulates ventilation and cardiovascular function in health and disease. European journal of heart failure 2021; 23(9): 1458–67.

44. Scott AC, Davies LC, Coats AJ, et al. Relationship of skeletal muscle metaboreceptors in the upper and lower limbs with the respiratory control in patients with heart failure. Clinical science (London, England : 1979) 2002; 102(1): 23-30.

45. García-Río F, Villamor A, Gómez-Mendieta A, et al. The progressive effects of ageing on chemosensitivity in healthy subjects. Respir Med 2007; 101(10): 2192–8.

46. Giannoni A, Gentile F, Buoncristiani F, et al. Chemoreflex and Baroreflex Sensitivity Hold a Strong Prognostic Value in Chronic Heart Failure. JACC Heart Fail 2022.

47. Narkiewicz K, Pesek CA, van de Borne PJ, et al. Enhanced sympathetic and ventilatory responses to central chemoreflex activation in heart failure. Circulation 1999; 100(3): 262–7.

48. Chua TP, Ponikowski P, Webb-Peploe K, et al. Clinical characteristics of chronic heart failure patients with an augmented peripheral chemoreflex. European heart journal 1997; 18(3): 480–6.

49. Dai L, Guo J, Hui X, et al. The potential interaction between chemosensitivity and the development of cardiovascular disease in obstructive sleep apnea. Sleep Med 2024; 114: 266–71.

50. Chowdhuri S, Pranathiageswaran S, Franco-Elizondo R, et al. Effect of age on long-term facilitation and chemosensitivity during NREM sleep. J Appl Physiol (1985) 2015; 119(10): 1088-96.

51. Paleczny B, Niewiński P, Rydlewska A, et al. Age-related reflex responses from peripheral and central chemoreceptors in healthy men. Clin Auton Res 2014; 24(6): 285–96.

52. Kato T, Kasai T, Suda S, et al. Prognostic effects of arterial carbon dioxide levels in patients hospitalized into the cardiac intensive care unit for acute heart failure. Eur Heart J Acute Cardiovasc Care 2021; 10(5): 497–502.

53. Naughton M, Benard D, Tam A, et al. Role of hyperventilation in the pathogenesis of central sleep apneas in patients with congestive heart failure. Am Rev Respir Dis 1993; 148(2): 330–8.

54. Lorenzi-Filho G, Azevedo ER, Parker JD, et al. Relationship of carbon dioxide tension in arterial blood to pulmonary wedge pressure in heart failure. The European respiratory journal 2002; 19(1): 37–40.

55. Carr J, Day TA, Ainslie PN, et al. The jugular venous-to-arterial PCO2 difference during rebreathing and end-tidal forcing: Relationship with cerebral perfusion. The Journal of physiology 2023; 601(19): 4251–62.

